# Deciphering the tissue-specific functional effect of Alzheimer risk SNPs with deep genome annotation

**DOI:** 10.1101/2023.10.23.23297399

**Authors:** Pradeep Varathan, Linhui Xie, Bing He, Andrew J. Saykin, Kwangsik Nho, Jingwen Yan

## Abstract

Alzheimer’s disease (AD) is a highly heritable brain dementia, along with substantial failure of cognitive function. Large-scale genome-wide association studies (GWAS) have led to a significant set of SNPs associated with AD and related traits. GWAS hits usually emerge as clusters where a lead SNP with the highest significance is surrounded by other less significant neighboring SNPs. Although functionality is not guaranteed with even the strongest associations in the GWAS, the lead SNPs have been historically the focus of the field, with the remaining associations inferred as redundant. Recent deep genome annotation tools enable the prediction of function from a segment of DNA sequence with significantly improved precision, which allows in-silico mutagenesis to interrogate the functional effect of SNP alleles. In this project, we explored the impact of top AD GWAS hits on the chromatin functions, and whether it will be altered by the genomic context (i.e., alleles of neighborhood SNPs). Our results showed that highly correlated SNPs in the same LD block could have distinct impact on the downstream functions. Although some GWAS lead SNPs showed dominating functional effect regardless of the neighborhood SNP alleles, several other ones do get enhanced loss or gain of function under certain genomic context, suggesting potential extra information hidden in the LD blocks.

## Introduction

Alzheimer’s disease (AD) is one of the most common form of brain dementia, along with which substantial failure of organs and mental issues arise. Nearly 10% of the US population above 65 have been accounted for AD in the US with the recent numbers projecting to 13.8 million by 2060 [1]. The heritability of AD is estimated to be between 60% and 80% [2]. Therefore, much work has been done in genetic association studies seeking the genetic architecture of AD since early 1990s, followed with several largescale genome-wide association studies (GWAS) and meta-analyses [3, 4, 5]. It is expected that these increasing findings will better delineate the pathways underlying disease. Yet, there remains a large gap of estimated heritability and that explained by existing GWAS findings [6, 7].

GWAS hits usually emerge as clusters where a lead SNP with the highest significance is surrounded by other less significant neighboring SNPs. This observation of hits in clusters aligns with the model of “haplotype blocks.” That is, genomic regions are inherited together as sets (i.e., haplotype blocks) and nearby variants within the blocks can be highly correlated, known as linkage disequilibrium (LD) [8, 9, 10]. Although functionality is not guaranteed with even the strongest associations in GWAS, the lead SNPs have been historically the focus of the field, with the remaining associations inferred as redundant [11]. In polygenic risk analysis where GWAS summary statistics was used to estimate the personal genetic risk of AD, the risk effect of neighboring SNPs are commonly excluded through pruning or clumping [12, 13]. Lead SNPs have also been widely used to assist with drug discovery since drug targets with genetic evidence of disease association are more likely to succeed [14]. Yet, lead SNPs identified from GWAS studies have not been consistent but rather nearby the same neighborhood [15]. Susceptibility locus in AD, reported as the nearest genes of lead SNPs, sometimes are also different even for the same SNP [15]. Taken together, information harbored in the neighborhood of lead SNPs may not be necessarily redundant. Focusing only on the lead SNPs will very likely limit our understanding of genetic factors in AD [11].

Recently, deep learning models have shown considerable promise in predicting the function of DNA sequence segment, such as transcription factor binding sites (TFBS) [16, 17]. These models attained high accuracy in predicting the underlying chromatin marks in a tissue-specific manner [18]. They also enable the in-silico mutagenesis in the input DNA sequence to interrogate the impact of each individual allele on the predicted function of input DNA sequence. That is, by changing one allele from major to minor in the input sequence, difference in the predicted function of sequence reflects the functional impact of candidate allele. In this paper, we will leverage a recent deep learning model, Expecto, and interrogate the downstream functional changes of top GWAS hits in AD [16, 17]. In particular, we aim to explore: 1) what are the functional impact of AD lead SNPs? 2) is there any difference in functional impact between lead SNPs and their neighboring ones in the same LD block?, and 3) whether the functional impact of AD lead SNPs will be affected by the genetic context (i.e., alleles of neighboring SNPs in the same input sequence)?

## Methods

### 0.1. GWAS candidate locus

AD risk SNPs were extracted from a large-scale genome wide association study (GWAS), the International Genomics of Alzheimer’s Project (IGAP). This study was performed with the imputed genotype of 11,480,632 single nucleotide polymorphisms (SNPs) from 21,982 Alzheimer’s disease cases and 41,944 cognitively normal controls. It is a combination of four consortia, including Alzheimer Disease Genetics Consortium (ADGC), European Alzheimer’s disease Initiative (EADI), Cohorts for Heart and Aging Research in Genomic Epidemiology Consortium (CHARGE), and Genetic and Environmental Risk in AD Consortium Genetic and Environmental Risk in AD/Defining Genetic, Polygenic and Environmental Risk for Alzheimer’s Disease Consortium (GERAD/PERADES) [19]. In this study, we focused on top 100 significant SNPs with smallest pvalue in IGAP. In addition, considering that top hits identified from GWAS studies have not been consistent but rather nearby the same neighborhood [15], neighboring genetic loci within the same linkage disequilibrium (LD) block of those top hits were also included. LD block information was estimated from 1000 Genome Project [20].

#### Deep genome annotation for allele-specific function

AD risk variants from GWASs are located predominantly in non-coding regions of the genome [21, 22, 23]. Therefore, gene regulation is speculated as part of the driving factors for Alzheimer’s disease. Recently, there has been a significant progress in predicting regulatory marks from raw DNA sequence using deep learning models [16, 17, 24, 25]. More specifically, these models can generate the likelihood of functions (e.g., DNase peak or binding of specific transcription factor) with a given DNA sequence segment. Allelespecific effect can be estimated by comparing the functional likelihood of two input sequences carrying major and minor allele respectively. For example, for DNase peak, if the likelihood generated from sequence with major allele is much higher than that from sequence with minor allele, it suggests a potential loss of DNase peak in minor allele carriers.

DeepSEA is a pre-trained deep genome annotation model built on the data from ENCODE and Roadmap Epigenomics projects [17, 16, 26, 27]. It takes as input a short DNA sequence centering around allele of interest and predicts its chromatin profiles, including transcription factor binding sites, histone marks, and DNase peaks across various tissues and cell types. In other words, it predicts whether any of those chromatin features exist in the input sequence. Given that majority of GWAS findings are from noncoding regions, these chromatin profiles could reveal the critical role of gene regulation underlying complex diseases. DeepSEA is trained to predict the likelihood of 919 chromatin features, and outperforms standard models, with median AUC as high as 0.958 across all chromatin features. The same group further extended DeepSEA to Expecto with 2002 chromatin features, on top of which we built our work.

#### Allele-specific function without genetic context

We first applied Expecto to evaluate the allele-specific function of AD GWAS candidate SNPs without considering the genetic context, i.e., all the neighboring SNPs in input sequence will have major alleles. Input sequence for Expecto is a 2000bp DNA sequence, centering around the SNP of interest. It was generated using Hg38 Genome assembly as the reference genome, which was used to train Expecto model. For each candidate SNP, two input sequences were generated: 1) one 2000bp reference sequence directly extracted from the reference genome, with 999bp upstream, 1000bp downstream and the center SNP taking major allele of European population. Neighboring SNPs in the flanking region, if any, all take major alleles. 2) another alternate sequence by replacing the center SNP with minor allele. By comparing the log odds of functional likelihood between reference and alternate sequences, we can estimate the downstream functional effect specific to AD risk SNPs and their neighboring ones (Fig. 1 (a)). Reversed reference and alternate sequences have also been examined but the predicted chromatin profiles are almost identical, so the results are not included. For both reference and alternate sequences, Expecto outputs functional likelihood of all chromatin features. Log odds change is derived by comparing the output log odds ratio to reflect the functional impact of minor allele [16].

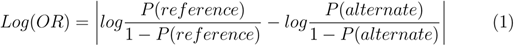

**Figure 1:**
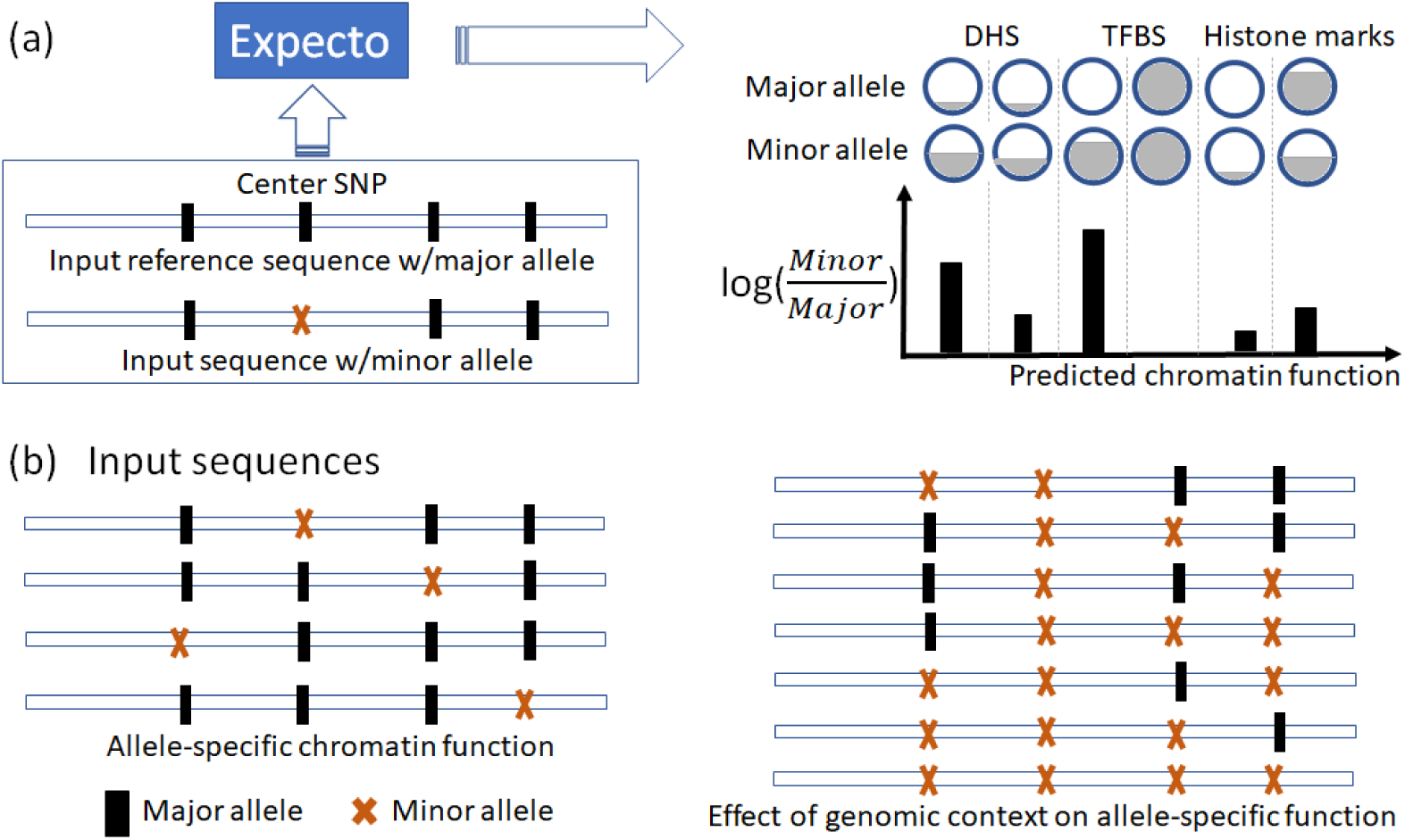
(a) Brief steps of Expecto to estimate the functional impact of the allele of interest (center SNP). Shadow inside the circle indicates the likelihood of one chromatin function given a specific input sequence. DHS: DNase I hypersensitive site, TFBS: Transcription factor binding site. (b) Input sequences used to estimate the allele-specific chromatin effect without genomic context (**Left**) and with genomic context (**Right**).

In this analysis, with a focus on Alzheimer’s disease, we manually screened all 2002 chromatin features in Expecto and included only 128 features highly relevant to brain (Table S1 in Supplementary Material). That is, these chromatin features are either from brain tissue or brain specific cell types like neuron, microglial and astrocyte. Monocyte is also included due to its close relationship with brain [28].

### 0.2. Allele-specific effect with influence of genetic context

Next, we tested the influence of neighboring SNPs on the allele-specific functional effect. That is, whether the genetic context provided by the neighboring SNPs located within 2000bp flanking region will change the functional impact of the center candidate SNP, e.g., enhancing or weakening the binding activity of certain transcription factor. Toward this, we generated a set of alternate sequences with in-silico mutagenesis, where the center SNP remains minor allele but neighboring SNPs selectively take minor alleles. We tested all possible combinations of major and minor alleles for neighboring SNPs, and examined whether any of the combinations will cause significant change in the functional effect of center SNP (Fig. 1 (b)). Finally, we used the ADNI genotype dataset to validate the effects of such combinations in AD, which was downloaded from the Alzheimer’s Disease Neuroimaging Initiative (ADNI) database (adni.loni.usc.edu).

#### E value generation

To evaluate the significance of our findings, we randomly selected 1,000,000 SNPs from chromosomes 1-22 and examined their allele-specific chromatin effect on all 128 brain-related chromatin features. As such, for each chromatin feature, we obtained a distribution of log odds ratio changes. On top of that, we estimated the empirical p-value of all log odds ratio values obtained using sequences around AD risk SNPs. Following [17], E-value is determined by the product of the log odds change (relative change) and the absolute change, and is formulated as below.

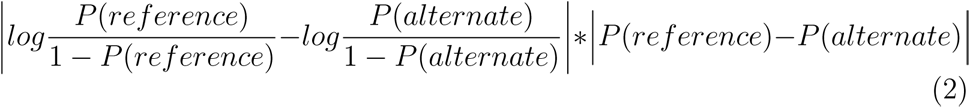

## Results

### Allele-specific effect without genetic context

After examining each individual AD risk SNPs and their neighborhood SNPs, we found 8 of them with noteworthy log odds ratio changes in brain-related chromatin features. Six are among top 100 AD GWAS candidate SNPs and two are in the 2000bp neighborhood of those top SNPs, with one as GWAS significant but not the other. Among the top AD GWAS SNPs, sequences with minor allele in rs157585 was predicted with prominent loss of function for DNase I hypersensitive sites (DHS) in gliblastoma cells, normal human astrocyte (NHA) and monocyte cells, and also histone marks in normal human astrocytes (Fig 2 (a)). Another top AD GWAS variant, rs74579864, leads to strong gain of function in acetylation of histones 2 and 3 at various positions in H1-Derived Neuronal Progenitor cells (NPC). rs35396326 is also associated with acetylation of histones 2 and 3 in H1-Derived Neuronal Progenitor cells with extremely high significance, but at different positions and in a negative way. Another prominent feature is the spike of the log odds ratio in gliobalstoma CTCF factor from rs75765623, located in the first intron of NECTIN2 gene. This variant is neither among the top GWAS SNPs nor a significant variant, but with highest log odds ratio change (empirical e-value = 0.0458). It is located only 70bp downstream of a significant AD GWAS hit rs12462573. Yet, chromatin effect associated with rs12462573 was minimal and negligible. When examined in the European population, we did not observe strong correlation between these two SNPs despite their closeness in physical location. It is therefore worth noticing that SNPs with significant p-value (i.e., lead SNPs) do not necessarily have the strongest functional impact in the brain. Actual functional impact could come from less significant variants located in the neighborhood of top hits.

**Figure 2:**
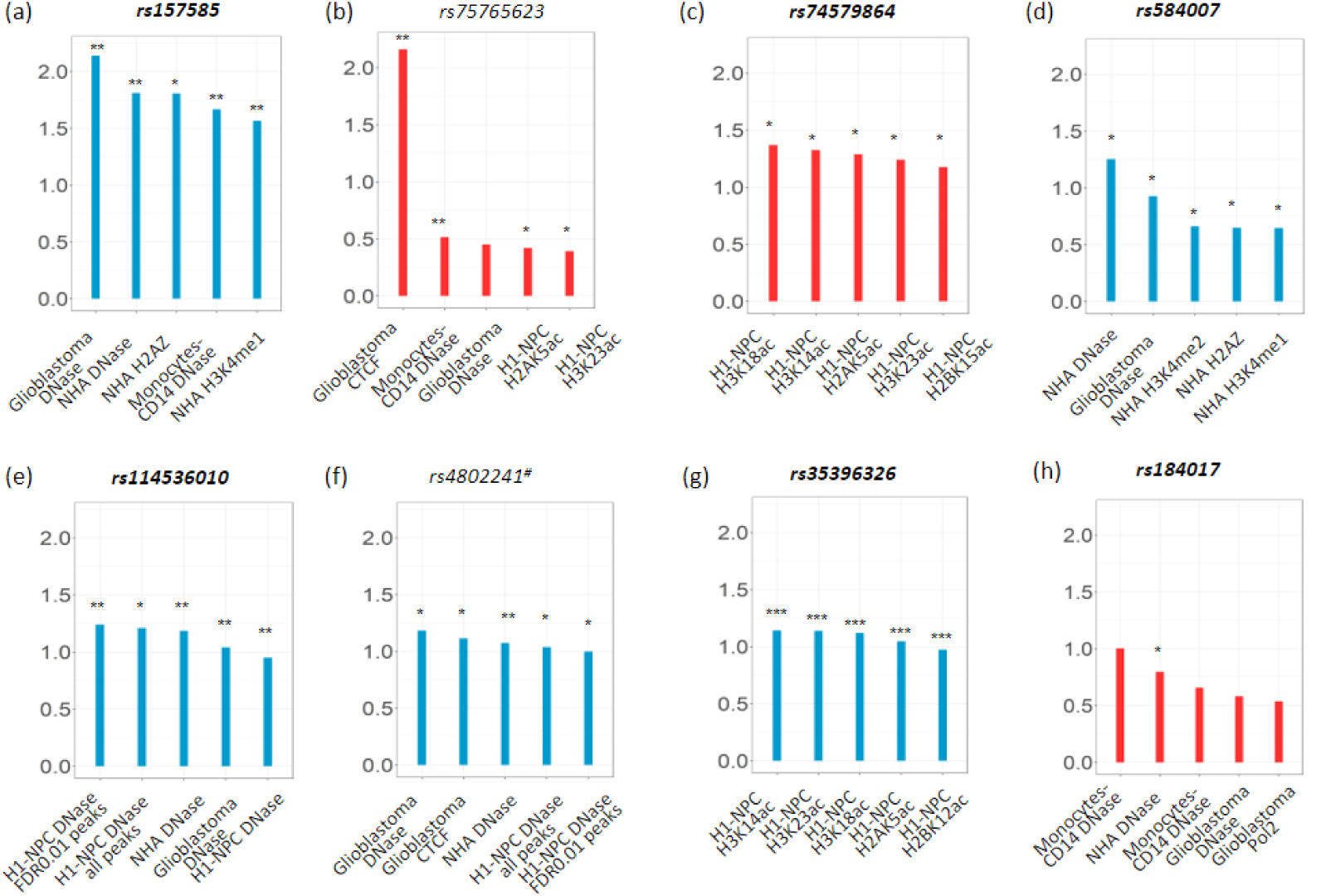
Top chromatin features affected by GWAS SNPs and their neighboring SNPs without genomic context. That is, each input sequence only have one SNP as minor allele. Only SNPs with at least one log odds change greater than 1 were included. Red is for positive log odds ratio change and blue for negative log odds ratio change, indicating gain and loss of function respectively. The variants in bold are among top 100 AD GWAS SNPs. Hash # indicates significant variants in the neighborhood of top GWAS SNPs while * indicates e-value *<* 5e-2, ** indicates e-value *<* 5e-3 and *** indicates e-value *<* 1e-6. NHA: Normal human astrocytes, H1-NPC: H1-derived neural progenitor cells.

### Highly correlated SNPs within LD block showed distinct allele-specific effect

Given that top 100 AD GWAS SNPs and their neighboring SNPs are close in physical locations, we further examined their linkage disequilibrium block structure using LDLink [29] (Supplementary Fig. 1) On top of that, 5 out of 8 SNPs with significant allele-specific effect, were found with highly correlated SNPs (corr *≥* 0.8) located in same LD block. Shown in Fig. 3 is the comparison of their allele-specific effect across highly correlated SNP groups. Each panel is a group of correlated SNPs in the same LD block and the first row is the SNP with significant allele-specific effect. Interestingly, these highly correlated variants seldom had similar predicted chromatin profiles. For example, rs74579864 (top GWAS hit) and rs4803761, rs4803762 are highly correlated (*R*^2^ = 0.956 and *R*^2^ = 0.978 respectively). However, minor allele of rs74579864 is associated with increased likelihood of histone marks in H1-derived neuronal progenitor cultured cells, but not those of rs4803761 and rs4803762. Similarly, sequences with minor allele of rs157585 showed strong negative effect in DNase hypertensive site in monocytes CD14 and glioblastoma cells, which we didn’t observe for its highly correlated neighbor rs157584 (*R*^2^ = 0.98). Taken together, our findings suggest that caution should be taken when pruning LD block to narrow down the number of SNPs for further investigation, which will likely result in loss of information.

**Figure 3:**
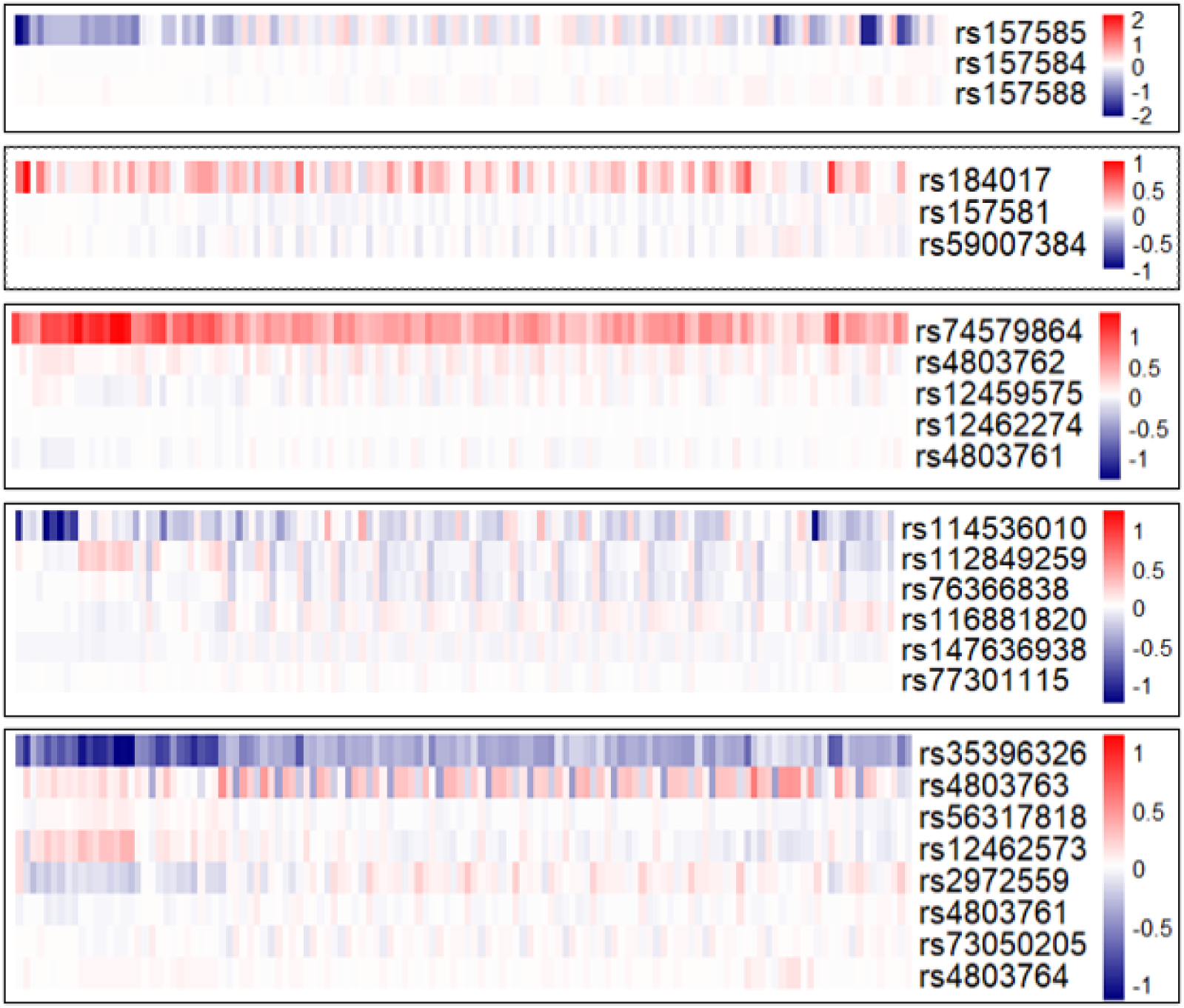
Highly correlated variants (R2 *≥* 0.8) of rs157585, rs184017, rs74579864, rs114536010 and rs35396326 from the same LD block show different functional effect.

### Allele-specific effect with genomic context

For all top 100 AD GWAS SNPs, we additionally examined the influence of genomic context on the allele-specific chromatin profiles. That is, whether the alleles of neighboring SNPs within 2000bp window will affect the function effect of the center SNP. As shown in Fig. 1 (b) right, input sequences centered around each SNP is modified by varying the allele of neighboring SNPs within the 2000bp window (i.e., major to minor allele). As such, we were able to identify 21 SNPs with strong effect on chromatin features (log odds change greater than 1, empirical *p ≤* 0.05). Predicted chromatin effect of these input sequences were compared with allele-specific effect without genomic context. Ultimately, four variants were observed with dominant effect, including rs157585, rs184017, rs114536010, and rs74579864. For each of these SNPs, input sequences carrying their minor allele have similar functional effect on chromatin features regardless of their genomic context in the 2000bp window. Three SNPs showed notable and significant log odds ratio change (*≥* 1, empirical *p ≤* 0.05), indicating the importance of genomic context for SNP annotation.

Variants rs1305062, rs2972559, and rs584007 showed notable and significant log odds ratio change (*≥* 1, empirical *p ≤* 0.05), indicating that their functional effect is dependent on the allele of neighboring SNPs. For sequences carrying minor allele of rs1305062, we observed significant loss of function for the CTCF binding sites in the glioblastoma cells, which becomes even worse when the input sequence also carries the minor allele of rs141864196 (Fig. 4 (b)). Similar effect was observed for rs2972559, which in combination with rs4802241 leads to more significant loss of function in the CTCF binding sites in the glioblastoma cells. Interestingly, the chromatin effect of rs2972559 (as a top GWAS hits) by itself is very weak (log odds change *< le* 0.5). On the other hand, minor allele in rs584007 is found associated with the loss of DNase I hypersensitive sites in normal human astrocyte derived cells. This loss of function becomes even more evident with the presence of minor allele in another SNP rs59325138, which has very weak chromatin effect by itself.

**Figure 4:**
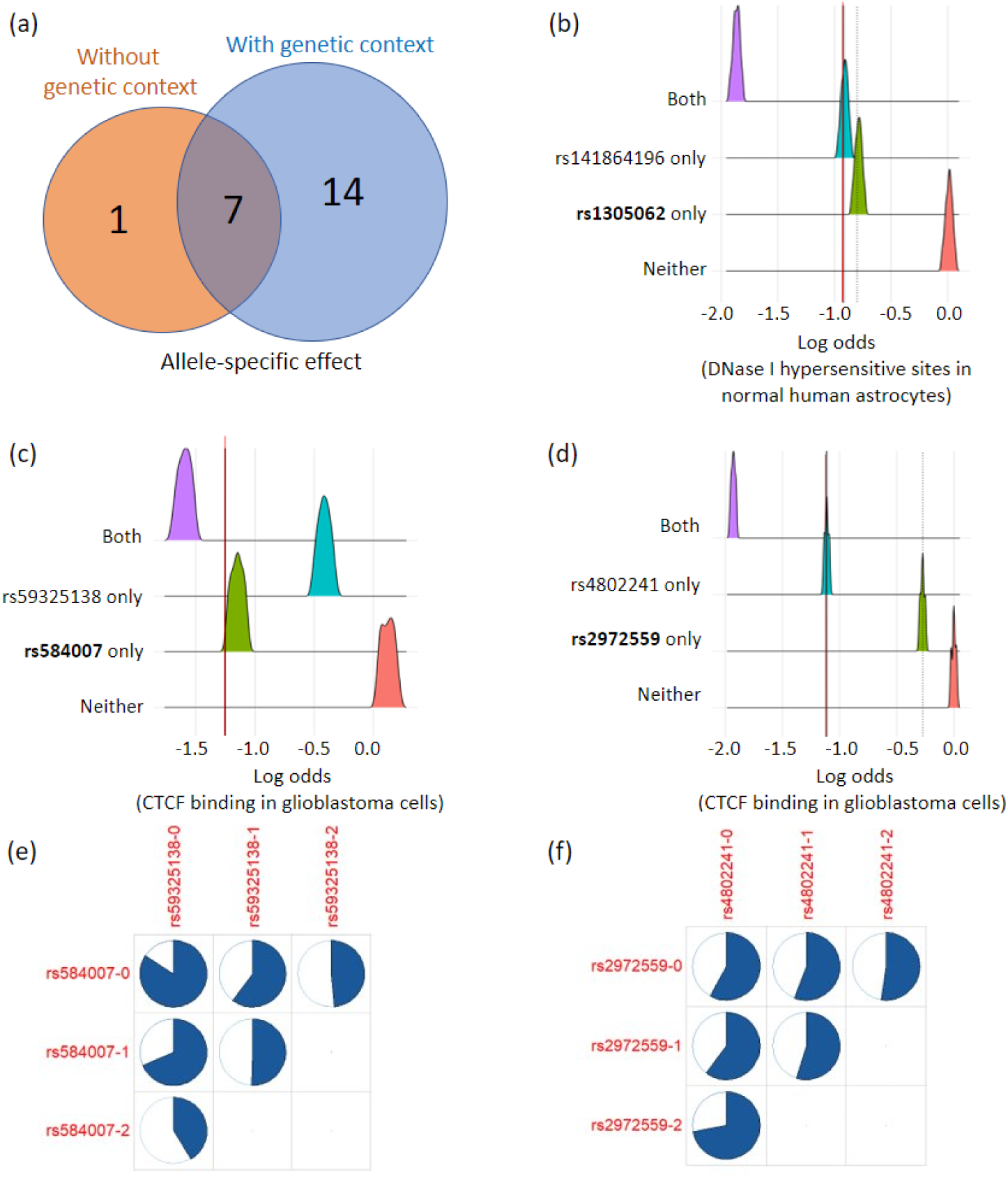
(a) Variants with significant predicted chromatin effect identified with and without genetic context (log odds ratio change *>* 1 and empirical *p ≤* 0.5). (b) Distribution of log odds change in CTCF binding in gliblastoma cells for input sequences centered around rs1305062. (c) Distribution of log odds change of DNase I hypersensitive sites in Normal Human Astrocytes for input sequences centered around rs584007. (d) Distribution of log odds change of CTCF binding in gliblastoma cells for input sequences centered around rs2972559 (e) Proportion of subjects developing AD in the ADNI cohort grouped by the genotype of rs584007/rs59325138. (f) Proportion of subjects developing AD in the ADNI cohort grouped by the genotype of rs2972559/rs4802241.

We further investigated the interaction effect of these three pairs of SNPs in the ADNI cohort [30]. Shown in Fig. 4 (e) and (f) are the distribution of subjects that ultimately developed AD during the follow-up visits. Row and columns indicate the genotype groups. Genotype groups without any ADNI subject are left blank. rs141864196 was not reported due to its missing genotype in ADNI. For both rs584007 (GWAS p = 1.056e-82, beta=-0.37) and rs59325138 (GWAS p = 6.945e-89, beta = −0.38), ratio of subjects developing AD decreases with the copies of minor allele, indicating their potential protective effect. Combined with their predicted chromatin effect, we speculate that deactivated DNase hypersensitive site in astrocyte cells may have a protective role in AD development. On the other hand, the number of minor allele in rs4802241 is not associated with AD development, where ratio of AD cases does not vary much across genotype groups. Its interactive partner rs2972559 (GWAS p = 3.448e-82) is related that more copies of minor allele confers higher risk of developing AD, but its predicted effect on CTCF binding is minimal. This mixed effect makes it difficult for us to conclude the effect of CTCF binding in glioblastoma on AD development, especially without support of carriers with 2 minor alleles in both SNPs.

## Discussion

This study investigated the chromatin function of sequences surrounding AD risk SNPs by leveraging the deep genome annotation tools. Amongst the top associated histone marks, most significant log odds changes are primarily observed in acetylation of histone H3, like H3K18ac and H3K23ac, all associated with GWAS SNP rs35396326 (beta = 0.38, p=1.35e-86 in GWAS). Acetylation levels of histones H3 and H4 has been previously reported to be overall lower in postmortem AD brains than in control brains. Among those, H3K18ac and H3K23ac were further validated as the most significantly hypoacetylated histone marks, along with H3K9ac, H3K27ac and H4K16ac [31]. In line with that, elevated levels of H3K14ac within the calpastatin promoter region is observed together with significantly decreased neuronal toxicity in neuroblastoma cells that underwent treatment to inhibit calcium-induced neuronal cell death [32]. While calcium-induced neuronal cell death is strongly associated with the pathophysiology of Alzheimer’s disease (AD), these evidence together suggests the potential neuroprotective role of histone acetylation. Our results provide extra support to this hypothesis that decreased histone acetylation is associated with the minor allele of AD risk SNP rs35396326 with positive beta coefficient (beta = 0.38, p=1.35e-86 in GWAS). In other words, minor allele in rs35396326 is associated with higher risk of developing AD and higher likelihood of decreased histone acetylation. Another group of chromatin features significantly associated with top AD GWAS hits are DNase in normal human astrocytes and monocytes. Most significant log odds change in DNase comes from *rs157585* and is specific to monocytes, astroytes and glioblastoma cells. In a DNase I footprinting analysis, mutations inside two DNAse hypersensitivite sites within recombinant *AP-2* were found associated with the regulation of the apoE promoter region, thereby implicating their role in the pathogenesis of AD [33]. Specifically, multiple DNase-I hypersenstivie sites were reported to be significantly associated to AD risk transcriptional factors in monocytes and macrophages [34]. The role of glial cells such as microglia, monocytes and astrocytes have been widely studied in neuroinflamation and AD, wherein the A*β* activated glial cells produce cytokines and chemokines which in turn activate pathways leading to demyelination, oxidative stress and eventually cell death [35]. While DNase I has been recently speculated as a potential therapeutic intervention for AD, cell-type specific DNase I activity is overall under explored in AD [36, 37].

Another two crucial findings of this investigation are that 1) SNPs in the same LD block with extremely high correlation (*≤* 0.9) sometimes have very distinct effect on the chromatin functions, and 2)variants not significant but in the neighborhood of GWAS hits could still have an impact on the downstream function (rs75762623 in Fig. **??** (b)). These results provided further proof that viewing LD-block as redundant information and having one variant as representative of the whole neighbourhood of LD block could significantly bias our understanding of the downstream function of GWAS findings.

In addition, we also observed significant genomic context effect on the functional effect of risk alleles. For the top 100 AD GWAS SNPs, carriers of minor alleles in two sets of neighboring SNPs were estimated with significantly more loss of function in CTCF binding activities in glioblastoma cells and DNase hypersentivitity sites in astrocytes. Such effect of genomic context does not necessarily comes from significant GWAS hits though. Even neighborhood SNPs that are not significant in the GWAS analysis, like rs4802241, could be of great importance as well to the further enhancement or decrease of downstream function. These findings provide evidence to support the importance of genetic context surrounding GWAS hits, which should not be simply treated as redundant information. Similar findings have only been recently reported in other diseases like Brugada syndrome [11]. While GWAS findings have been increasingly leveraged for many important downstream applications like polygenic risk estimation and discovery of drug targets, caution should be taken when interpreting the GWAS hits, especially considering the limited replicability of polygenic risk scores and failure of many clinical trials. Taken together, our results suggest the need for the re-analysis of published AD GWAS data and reconsideration of future application plans of GWAS findings.

This work has several limitations though that merit further consideration. First, given that long input sequence and large number of variants could lead to exponentially high number of genotype combinations as genomic context, we constrained this project to top 100 AD GWAS SNPs, which are mostly located around APOE region and employed Expecto with 2000bp input sequence. Moreover, we refrained from using haplotypes from 1000 genome project due to that they are mostly healthy participants. As such, we examined all possible combinations of minor alleles across the SNPs within 2000bp input window, some of which may not exist in population data. All of these limitations could be addressed with further haplotype estimation from phased genotype in large cohorts like UKBiobank or ADNI. Overall, this study provided a new perspective of interpreting the GWAS findings and new evidence to support the non-redundancy hypothesis of neighborhood variants surrounding GWAS hits. More in-depth work warrants further effort to investigate the functional effect of GWAS hits as clusters.

## Supporting information

All supplementary materials

## Data Availability

All data produced in the present study are available upon reasonable request to the authors and are also available online

https://adni.loni.usc.edu/new-admc-metabolomics-data-is-available/

https://www.ukbiobank.ac.uk/

## Acknowledgements

Data collection and sharing for this project was funded by the Alzheimer’s Disease Neuroimaging Initiative (ADNI) (National Institutes of Health Grant U01 AG024904) and DOD ADNI (Department of Defense award number W81XWH-12-2-0012). ADNI is funded by the National Institute on Aging, the National Institute of Biomedical Imaging and Bioengineering, and through generous contributions from the following: AbbVie, Alzheimer’s Association; Alzheimer’s Drug Discovery Foundation; Araclon Biotech; BioClinica, Inc.; Biogen; Bristol-Myers Squibb Company; CereSpir, Inc.; Cogstate; Eisai Inc.; Elan Pharmaceuticals, Inc.; Eli Lilly and Company; EuroImmun; F. Hoffmann-La Roche Ltd and its affiliated company Genentech, Inc.; Fujirebio; GE Healthcare; IXICO Ltd.; Janssen Alzheimer Immunotherapy Research & Development, LLC.; Johnson & Johnson Pharmaceutical Research & Development LLC.; Lumosity; Lundbeck; Merck & Co., Inc.; Meso Scale Diagnostics, LLC.; NeuroRx Research; Neurotrack Technologies; Novartis Pharmaceuticals Corporation; Pfizer Inc.; Piramal Imaging; Servier; Takeda Pharmaceutical Company; and Transition Therapeutics. The Canadian Institutes of Health Research is providing funds to support ADNI clinical sites in Canada. Private sector contributions are facilitated by the Foundation for the National Institutes of Health (www.fnih.org). The grantee organization is the Northern California Institute for Research and Education, and the study is coordinated by the Alzheimer’s Therapeutic Research Institute at the University of Southern California. ADNI data are disseminated by the Laboratory for Neuro Imaging at the University of Southern California.

