## Supplementary material for "Deciphering the tissue-specific functional effect of Alzheimer risk SNPs with deep genome annotation": All supplementary materials

### Slide 1
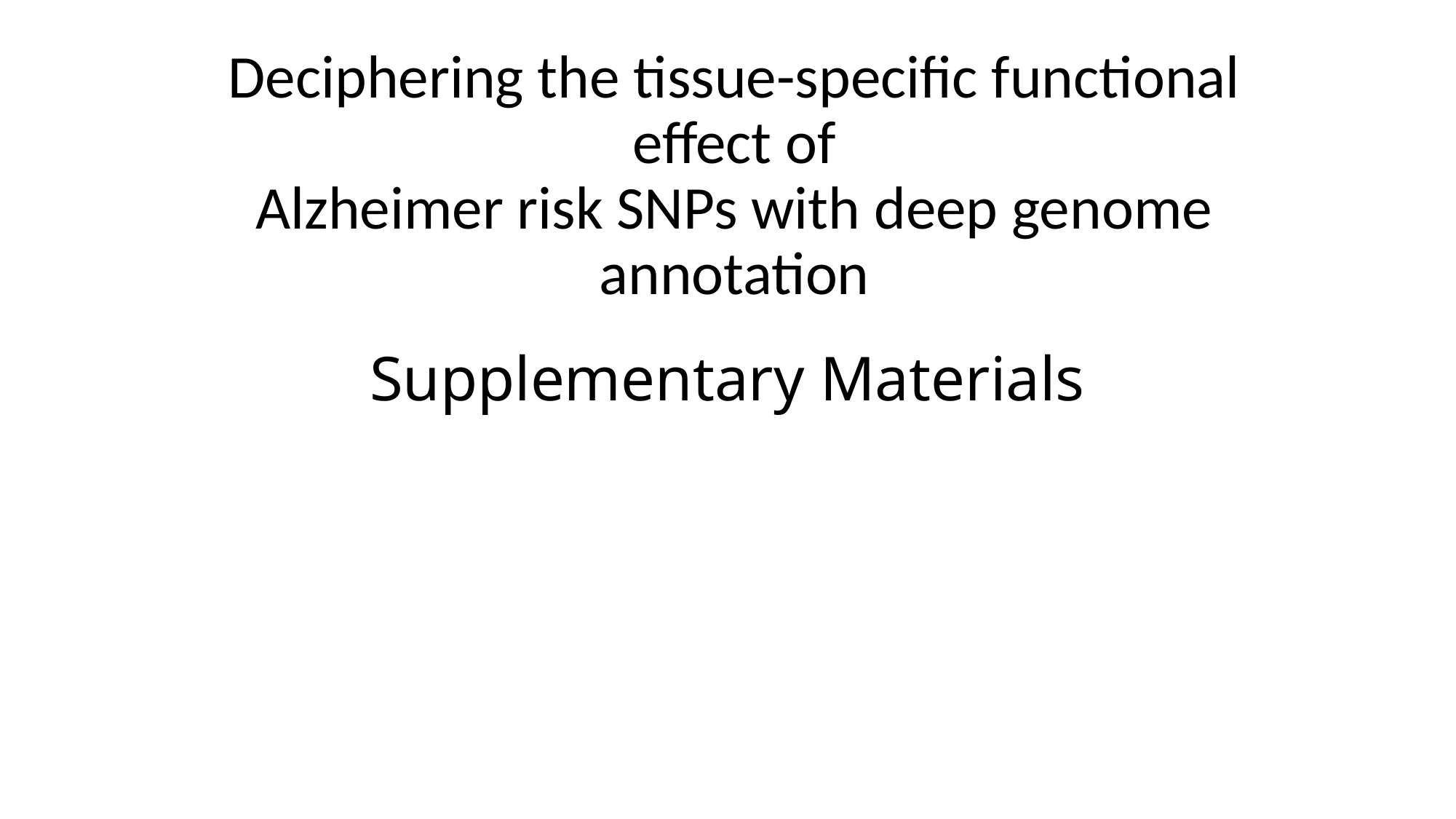

Deciphering the tissue-specific functional effect of
Alzheimer risk SNPs with deep genome annotation
#### Supplementary Materials

### Slide 2
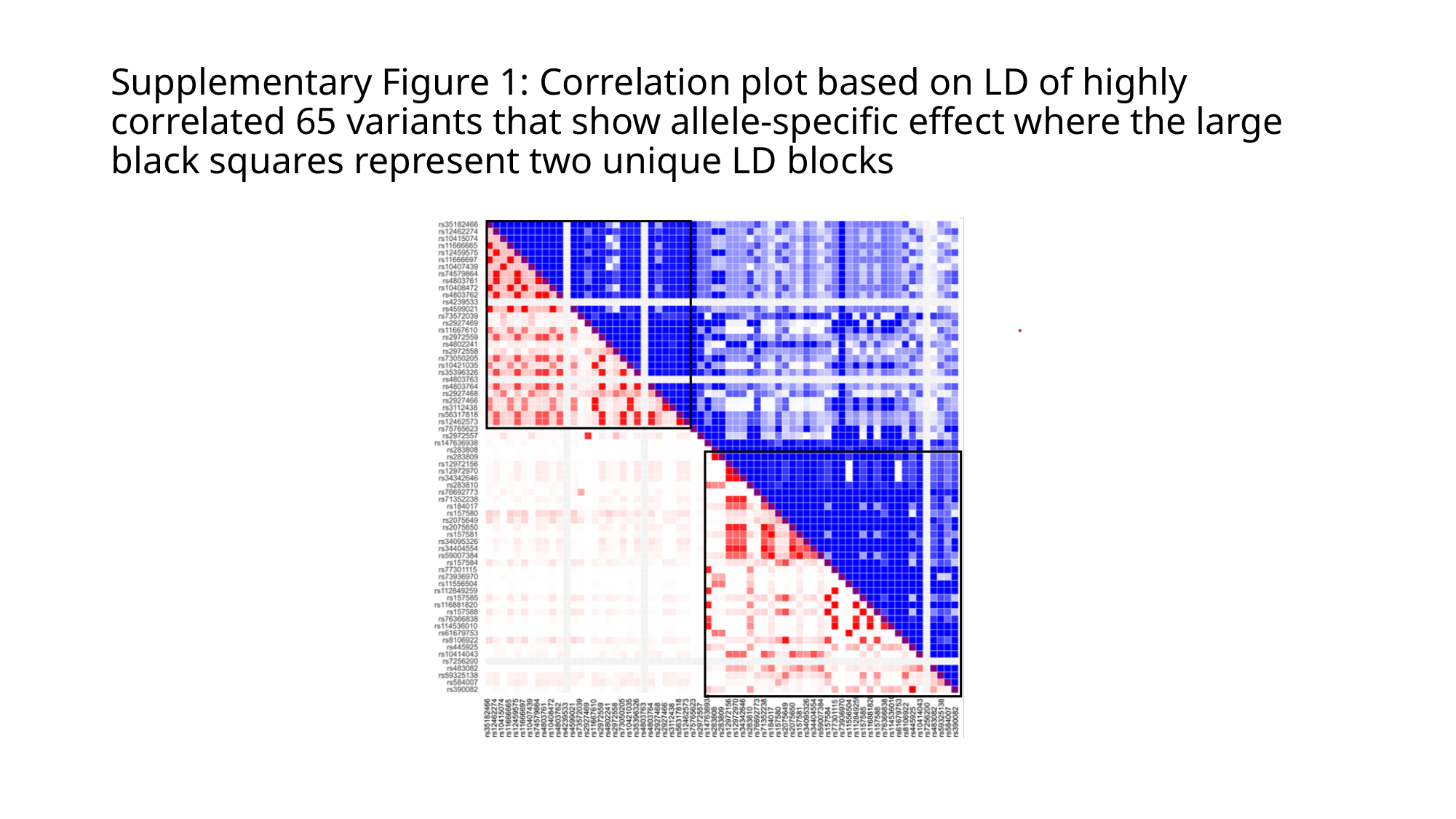

#### Supplementary Figure 1: Correlation plot based on LD of highly correlated 65 variants that show allele-specific effect where the large black squares represent two unique LD blocks

### Slide 3
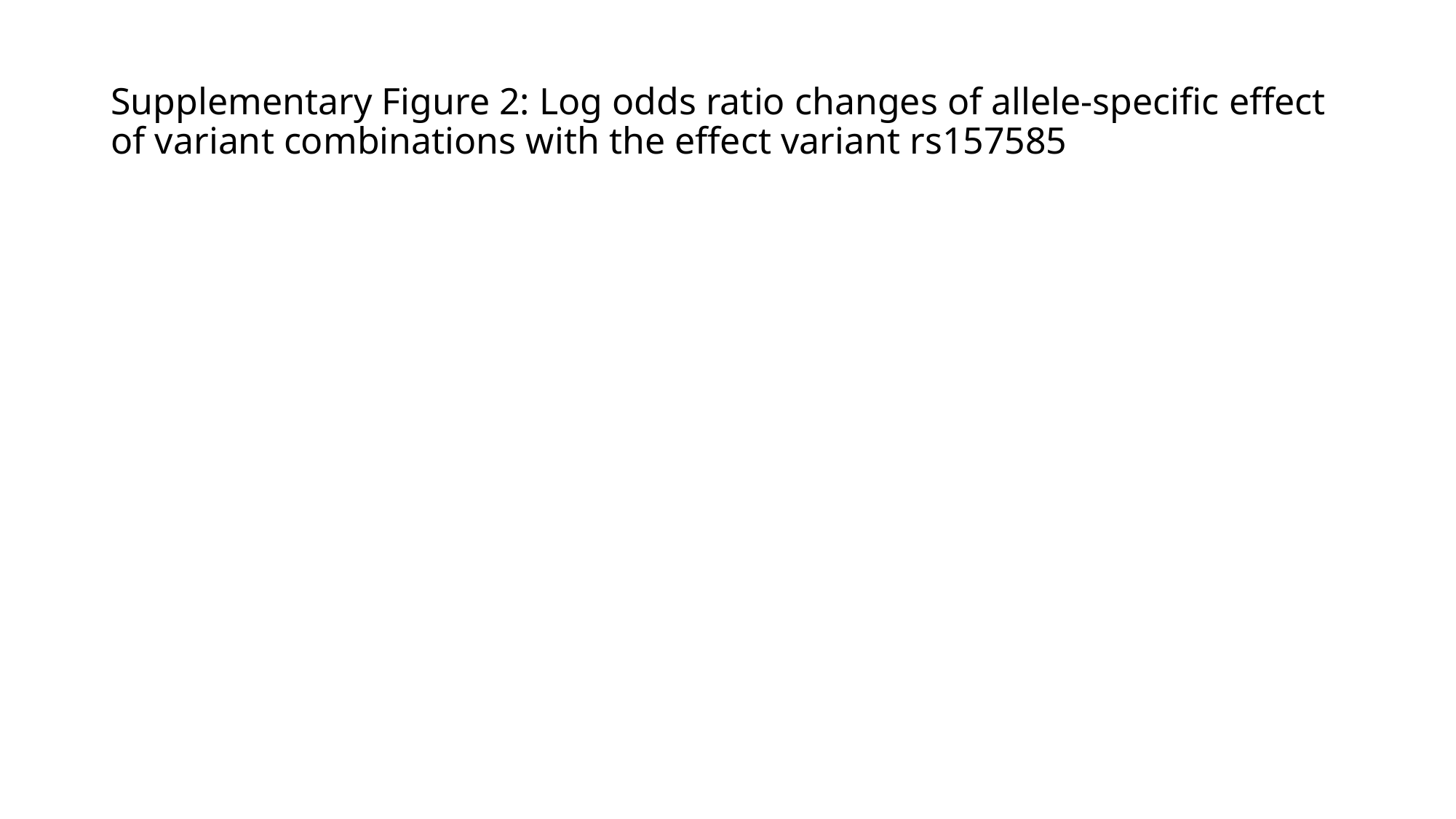

#### Supplementary Figure 2: Log odds ratio changes of allele-specific effect of variant combinations with the effect variant rs157585

### Slide 4
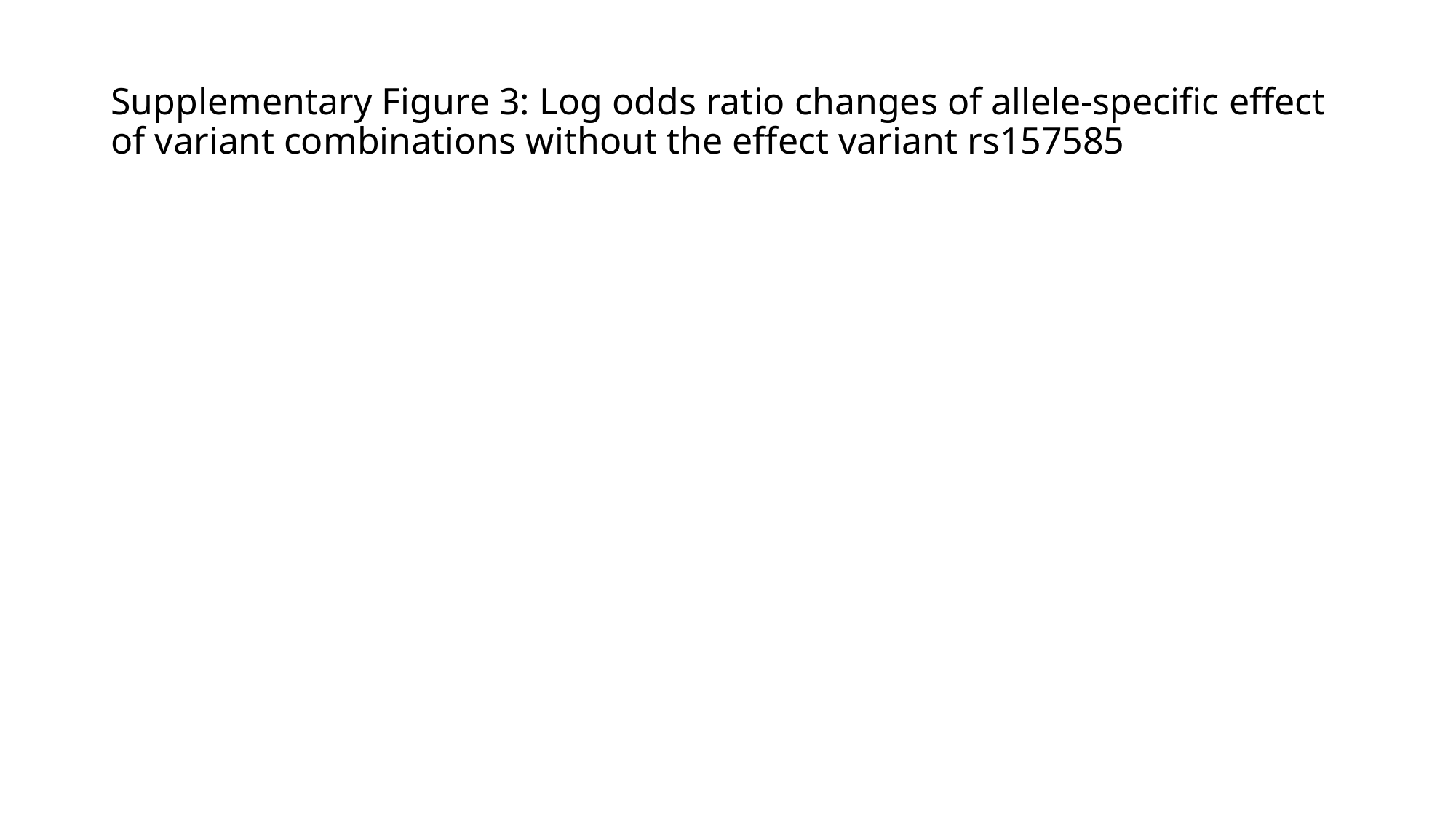

#### Supplementary Figure 3: Log odds ratio changes of allele-specific effect of variant combinations without the effect variant rs157585

### Slide 5
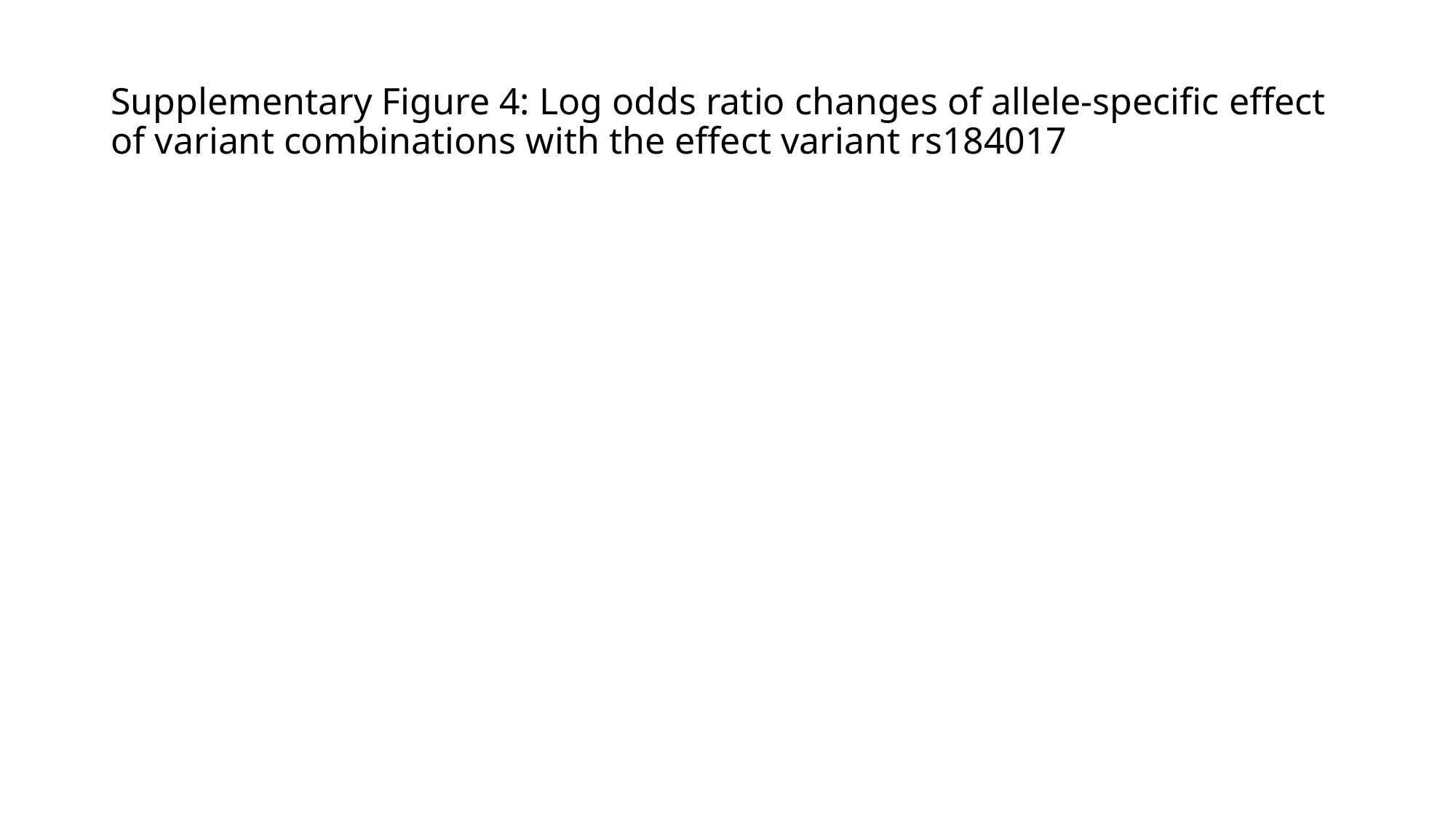

#### Supplementary Figure 4: Log odds ratio changes of allele-specific effect of variant combinations with the effect variant rs184017

### Slide 6
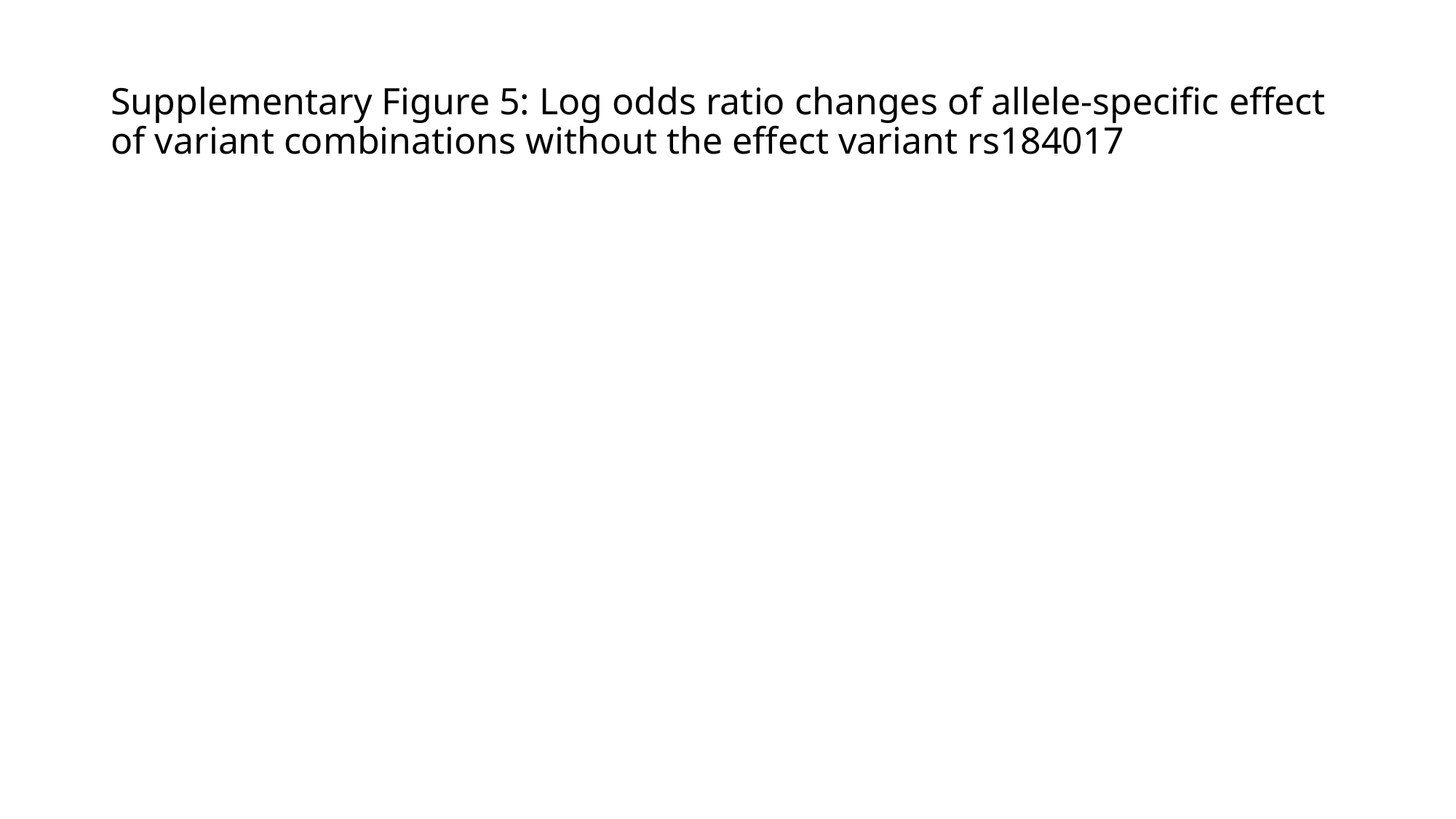

#### Supplementary Figure 5: Log odds ratio changes of allele-specific effect of variant combinations without the effect variant rs184017

### Slide 7
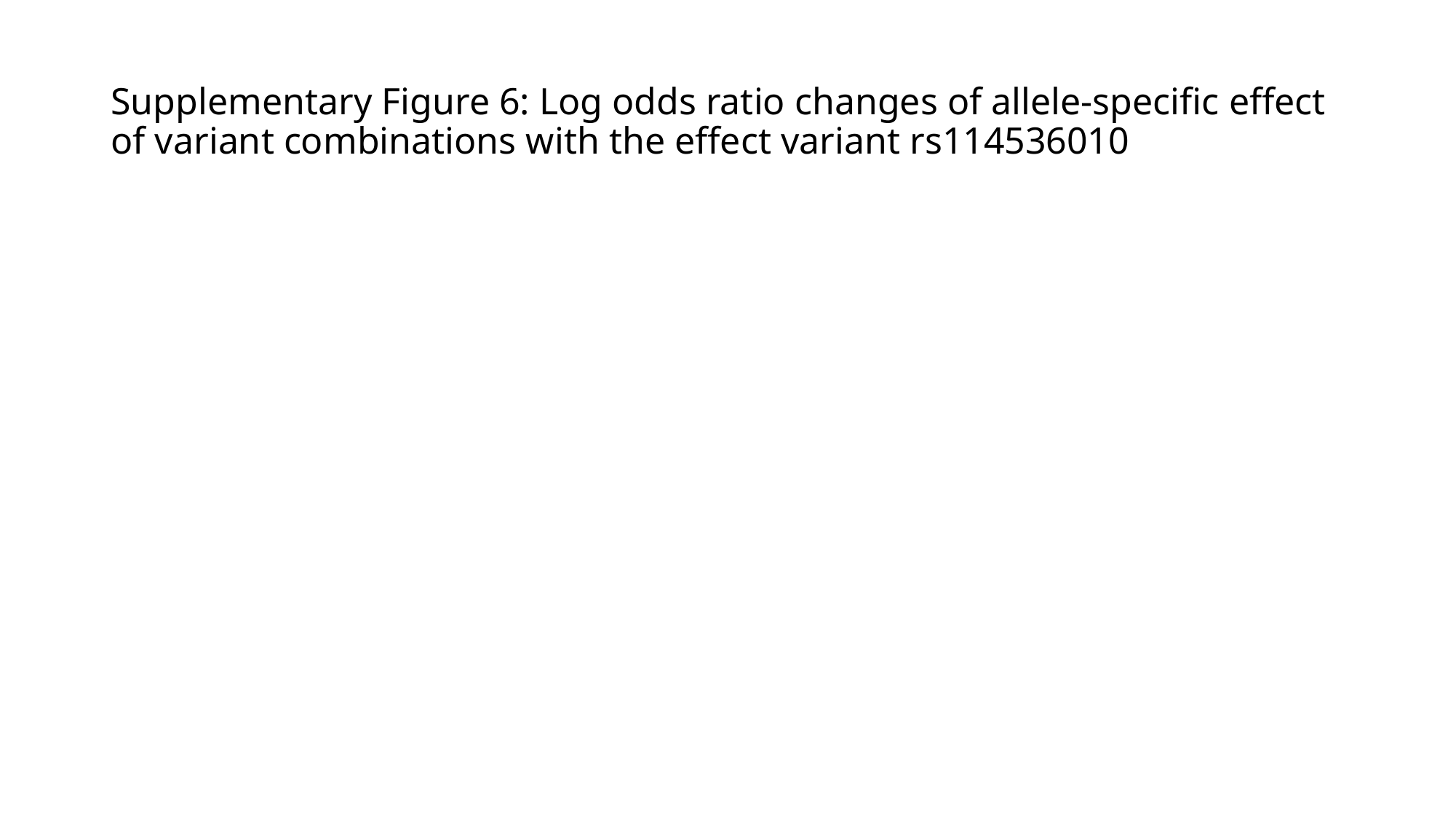

#### Supplementary Figure 6: Log odds ratio changes of allele-specific effect of variant combinations with the effect variant rs114536010

### Slide 8
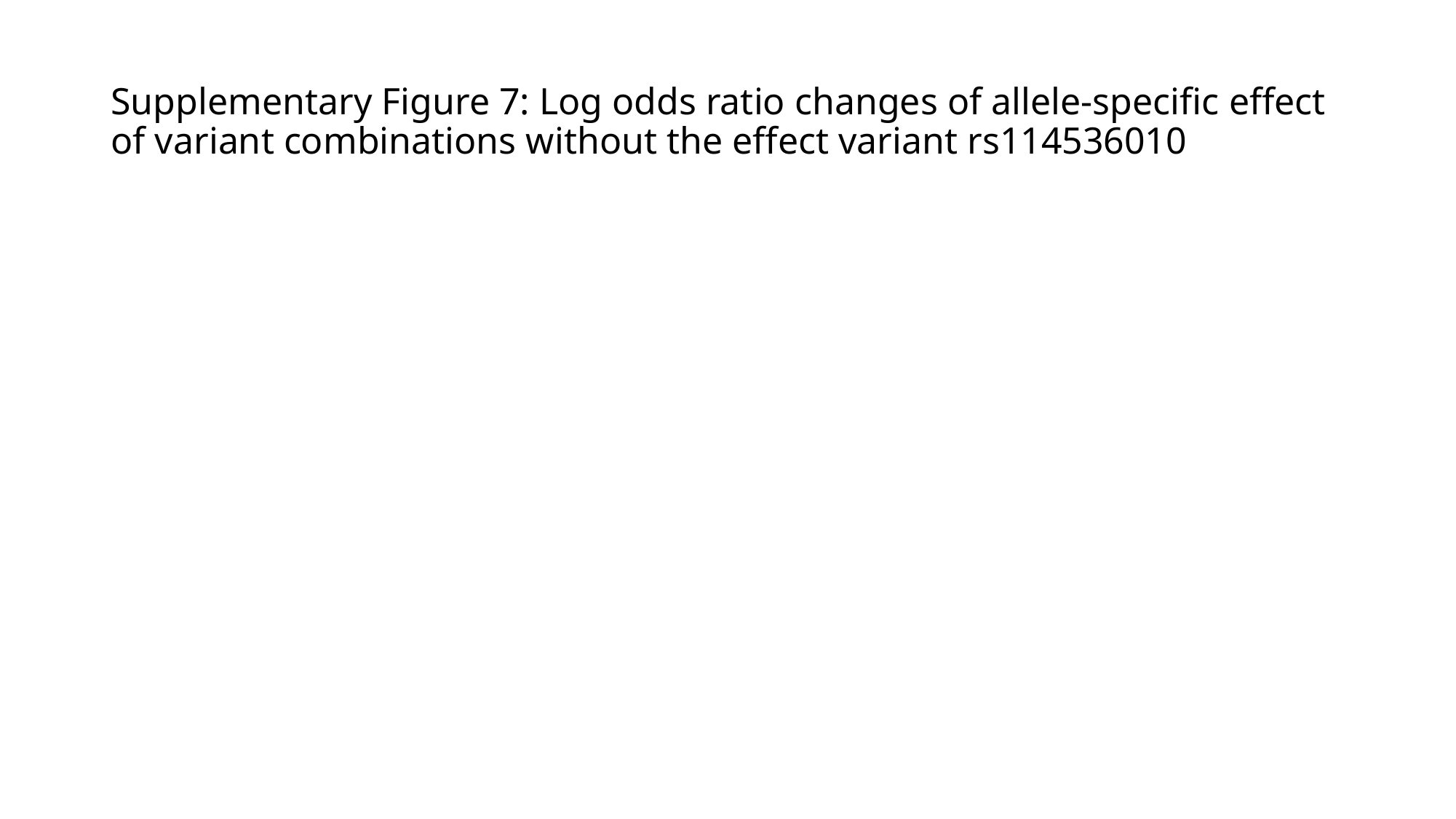

#### Supplementary Figure 7: Log odds ratio changes of allele-specific effect of variant combinations without the effect variant rs114536010

### Slide 9
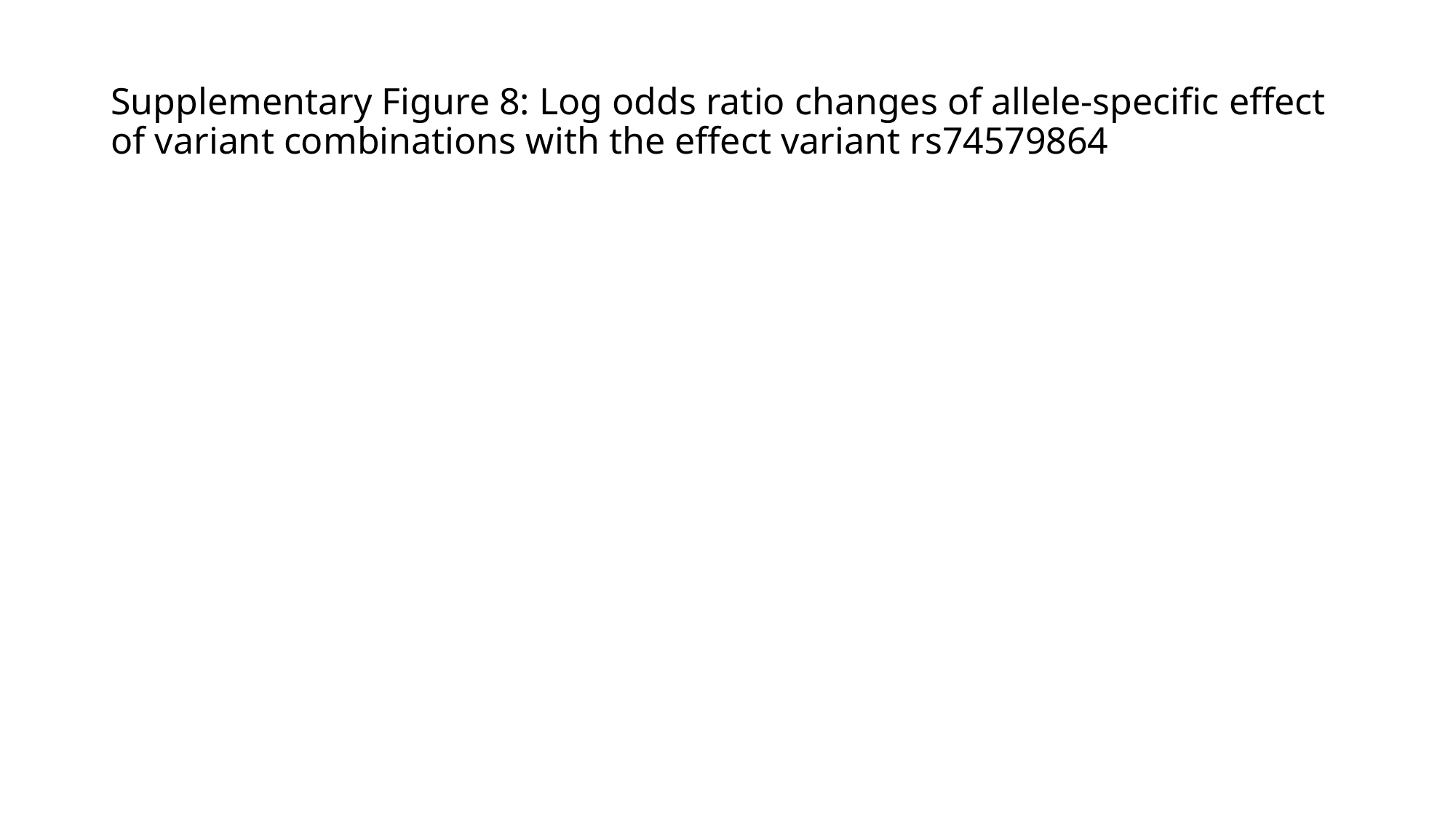

#### Supplementary Figure 8: Log odds ratio changes of allele-specific effect of variant combinations with the effect variant rs74579864

### Slide 10
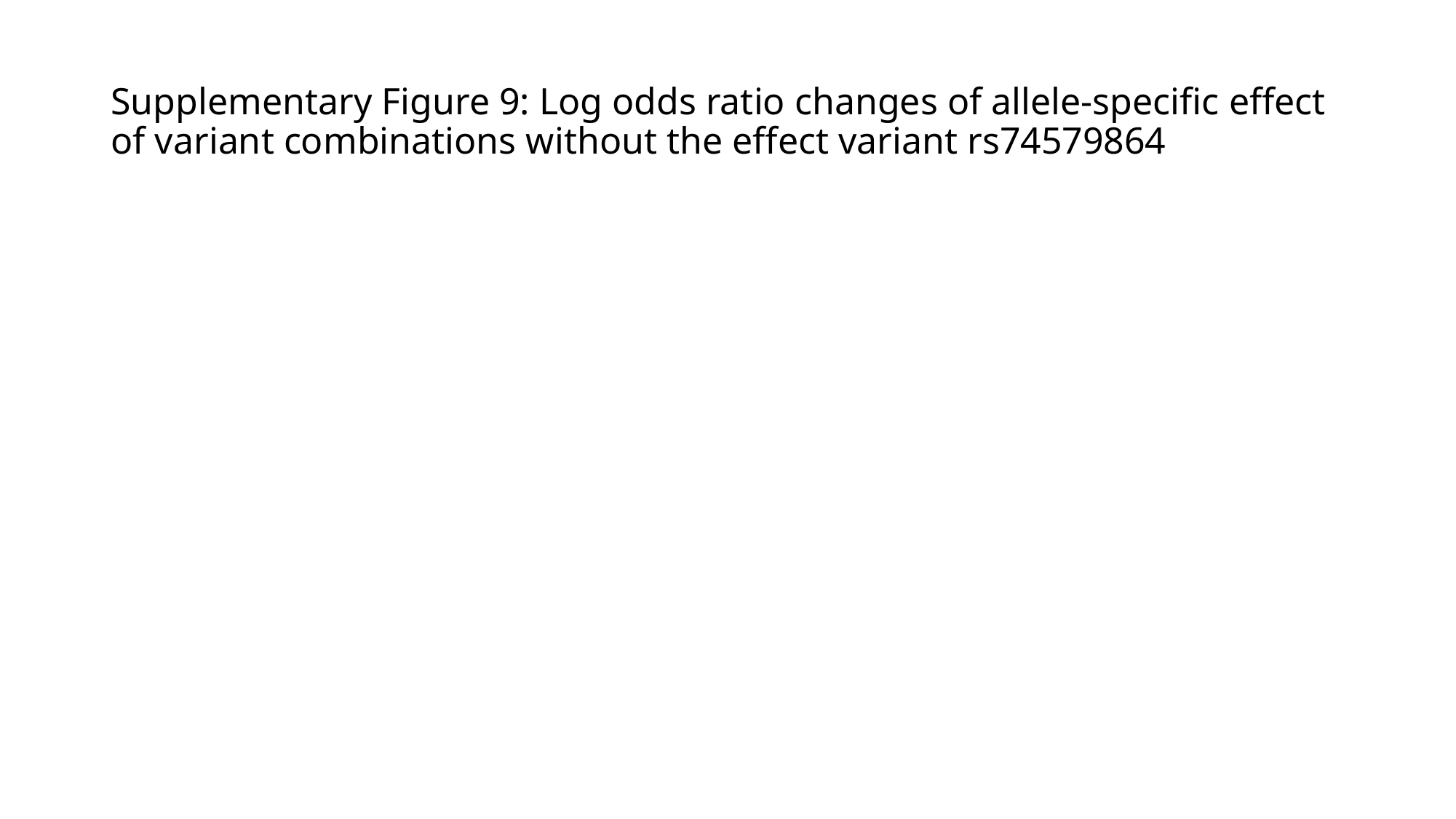

#### Supplementary Figure 9: Log odds ratio changes of allele-specific effect of variant combinations without the effect variant rs74579864

### Slide 11
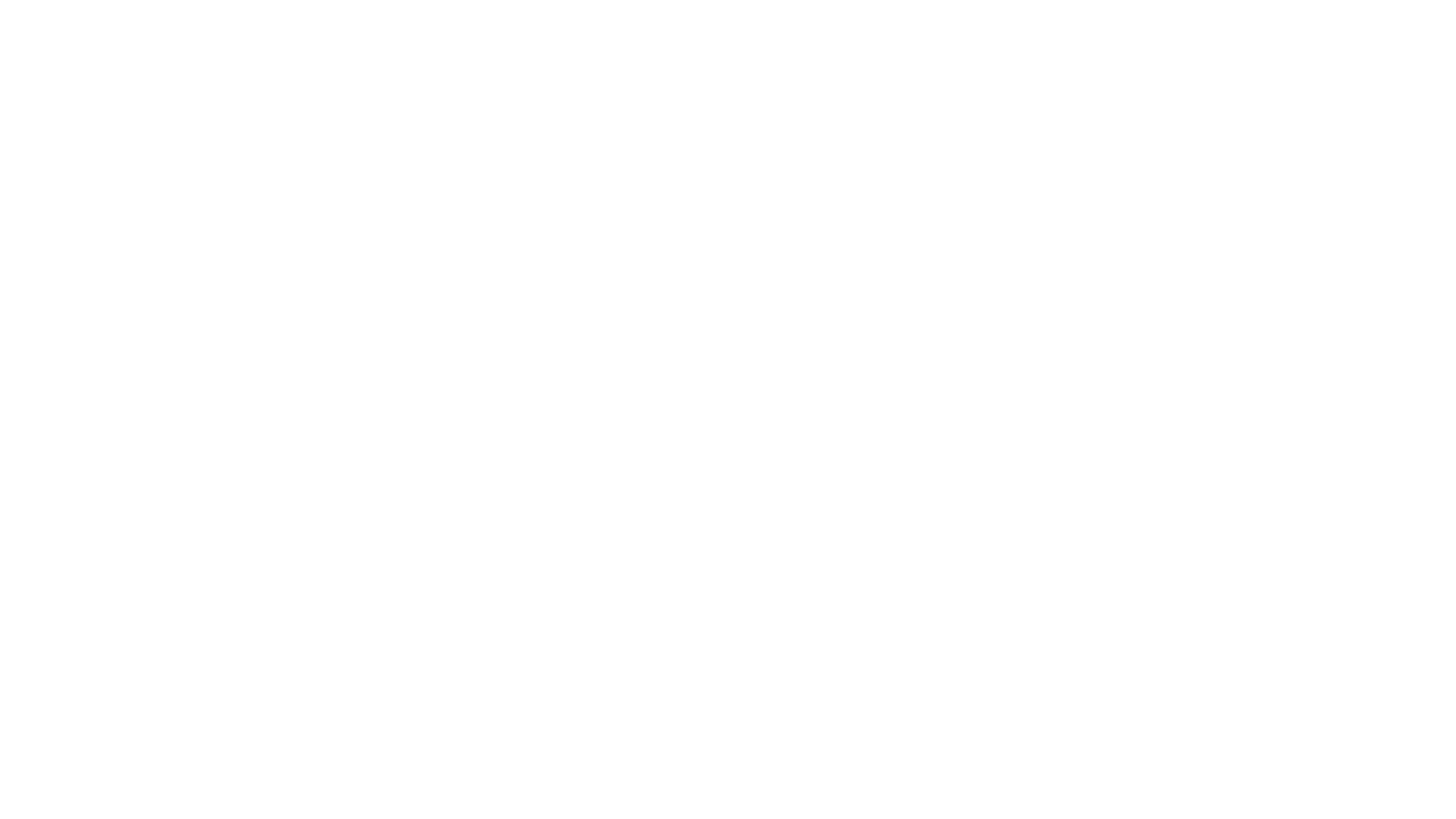

#
